# Improving ALS Detection and Cognitive Impairment Stratification with Attention-Enhanced Deep Learning Models

**DOI:** 10.1101/2024.09.22.24313406

**Authors:** Yuqing Xia, Jenna M Gregory, Fergal M Waldron, Holly Spence, Marta Vallejo

## Abstract

Amyotrophic lateral sclerosis (ALS) is a fatal neurological disease marked by motor deterioration and cognitive decline. Early diagnosis is challenging due to the complexity of sporadic ALS and the lack of a defined risk population. In this study, we developed Miniset-DenseSENet, a convolutional neural network combining DenseNet121 with a Squeeze-and-Excitation attention mechanism, using 190 autopsy brain images from the Gregory Laboratory at the University of Aberdeen. The model distinguishes ALS patients from controls with 97.37% accuracy and detects cognitive impairments, a critical but underdiagnosed feature of ALS. Miniset-DenseSENet outperformed other transfer learning models, achieving a sensitivity of 1 and specificity of 0.95. These findings suggest that integrating transfer learning and attention mechanisms into neuroimaging can enhance diagnostic accuracy, enabling earlier ALS detection and improving patient stratification. This model has the potential to guide clinical decisions and support personalized therapeutic strategies.

## 1 Main

Amyotrophic lateral sclerosis (ALS) is a condition that progressively degenerates the neurons responsible for motor control [1]. As a patient’s motor neurons degenerate and die, the patient’s brain loses the ability to initiate and control voluntary movement and eventually dies from respiratory failure [2]. ALS has a median survival of only 2–4 years. The ALS Therapy Development Institute projects a global rise in ALS instances from 222,801 in 2015 to 376,674 by 2040, marking a 69% surge [1]. Moreover, the average delay from the first appearance of symptoms to the final diagnosis is about 14 months [3].

In an analysis of autopsy cases from ALS patients, a specific protein called the TAR DNA-binding protein 43 (TDP-43) was found to accumulate abnormally in the cytoplasm [4]. This abnormal accumulation is observed in the motor neurons of nearly all patients with sporadic ALS and in the majority of patients with familial ALS (*>* 84%) [5]. The frequent presence of TDP-43 in almost all sALS patients and most fALS patients suggests that it may play a key role in the ALS disease process [2]. Additionally, Gregory *et al.* found that although all ALS patients exhibit TDP-43 pathology in extramotor brain regions, only some present with cognitive impairment [4]. The progression and ultimate outcome of ALS vary significantly between patients, making it difficult to accurately predict the disease’s course [6, 7].

Deep learning models have achieved unexpected progress in classification and recognition tasks [8], given their ability to recognize complex and subtle patterns and relationships. However, capturing complex disease features in small datasets is challenging [9], [10]. Therefore, developing robust models that can effectively handle small datasets and capture complex disease characteristics remains an important research direction. Data augmentation and transfer learning are two widely adopted strategies to alleviate the limitations of small datasets [11].

Data augmentation involves creating new training samples by applying various transformations to the original data [12]. Data augmentation methods involve rotating or reflecting the original image, scaling it up or down, and translating it to create new images. Then, the model can become more robust to changes in image orientation and position, ultimately enhancing its generalization ability [13].

Transfer learning is another effective method to address the issue of limited training data. Using models pre-trained on large datasets such as ImageNet allows transfer learning to transfer the learned features to a new, smaller dataset [14]. This process involves fine-tuning the pre-trained model on a specific medical dataset, which can improve the model’s initial performance and reduce the need for a large amount of labeled data [15].

However, data augmentation and transfer learning alone may not fully address the complexity of disease features in medical images. To further enhance model performance, attention mechanisms to optimize feature extraction and highlight the most relevant parts of the input data. Attention mechanisms can be intuitively explained using the human visual system [16], and they have good integration properties with many methods being very lightweight, such as the squeeze-and-excitation (SE) model and the convolutional block attention module (CBAM) model. This makes them well-suited for combination with deep learning [17]. Generally, attention mechanisms determine which information is important by assigning weights. This process improves model accuracy and provides better interpretability of the results. Specifically, the SE module’s channel-level attention mechanism can better capture and utilize finegrained features within DenseNet, enhancing the model’s recognition and classification performance.

In this study, we exploit the combination of transfer learning and attention mechanism to develop an effective model framework named Miniset-DenseSENet. The model leverages the powerful feature extraction capabilities of pre-trained DenseNet121 and combines SE modules to enhance feature representation. By focusing on the most relevant features in the dataset, our approach aims to improve the diagnosis and understanding of ALS.

## 2 Results

### 2.1 Original Image Quality Analysis

The inspection of the dataset shown in Figure 1 found that the mean distribution of contrast and brightness is relatively concentrated, and there are no extreme outliers, indicating that most images are relatively consistent in brightness and contrast. The SNR distribution is narrow, which indicates that the images are relatively balanced in signal quality without excessive noise. However, the distribution of the Laplacian variance shows that there are varying degrees of image sharpness but no outliers. The images with the minimum and maximum Laplacian variance are shown in Figure 1(1) and Figure 1(2). It can be observed that most images are acceptable in terms of clarity. These distribution plots indicate that the image quality of the dataset is appropriate and can be used for further data enhancement and deep learning training. See A.1 for further details.

**Fig. 1:**
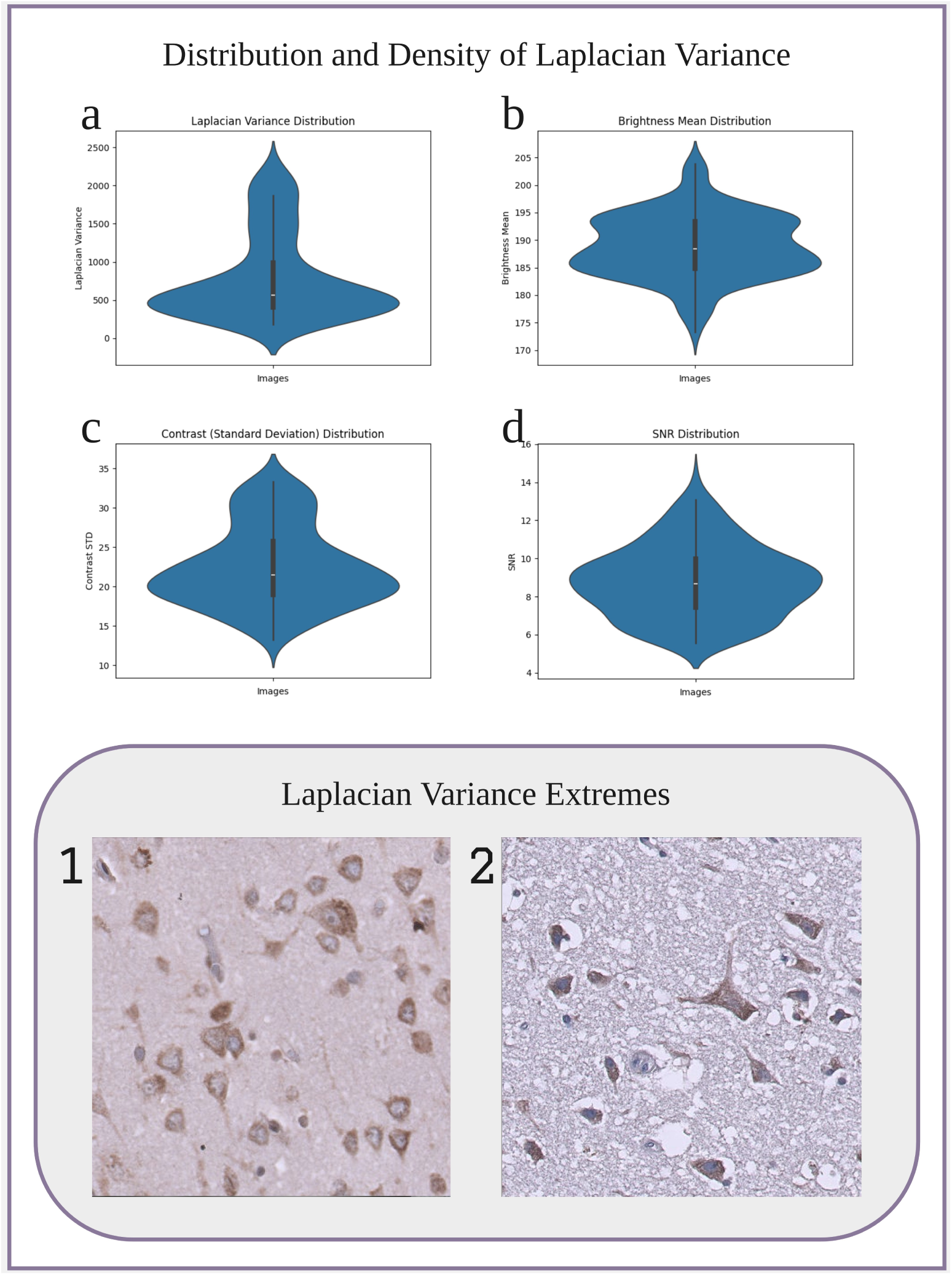
Top: Violin plots showing the distribution and density of four image features: (a) Laplacian variance, (b) signal-to-noise ratio (SNR), (c) brightness mean, and (d) contrast. These plots illustrate the variability and spread of these metrics across the dataset. Bottom: Two ALS image samples representing the extremes of Laplacian variance. Image 1 shows the minimum variance, while Image 2 shows the maximum variance, highlighting the visual differences in image sharpness between the two samples. 4

### 2.2 Performance Evaluation over Different Models

We undertook a detailed training and evaluation of Miniset-DenseSENet and four other distinct models: ResNet18, DenseNet121, ResNet18+SE, and ResNet18+CBAM. Figure 2(top) shows the accuracy convergence as the number of training iterations increases in all the models considered. Figure 2(middle) shows accuracy and MCC median values among the five runs. A comparative analysis of these figures reveals that the accuracy of the five transfer learning models is significantly better than that of the conventional CNN approach. Notably, the accuracy of the DenseNet model augmented with the SE module consistently exceeds 90%, with an optimal performance of 97.37%.

**Fig. 2:**
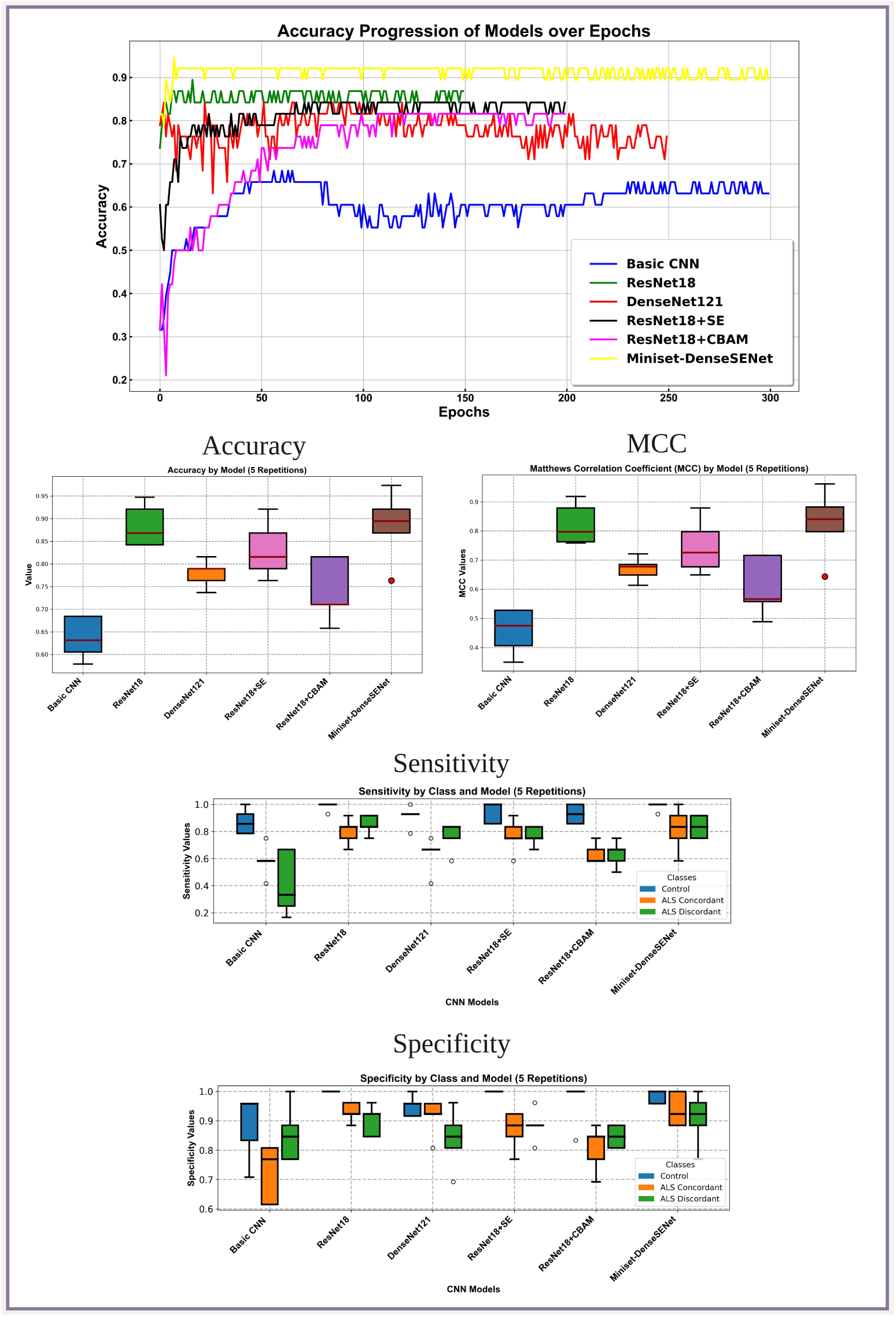
The figure illustrates the performance comparison between different models, arranged from top to bottom. At the top, the graph displays the accuracy values collected over consecutive epochs for six5 models: Miniset-DenseSENet, ResNet18, ResNet18+SE, ResNet18+CBAM, DenseNet121, and Basic CNN. The models are listed in order of performance from best to worst accuracy convergence. In the middle row, boxplots show the accuracy and Matthews correlation coefficient (MCC) over five repetitions, where Miniset-DenseSENet demonstrates the highest accuracy and MCC, followed by ResNet18. The final row presents sensitivity and specificity values for each model, with results and variance aligning with the accuracy and MCC observations from previous rows. These plots confirm the superior performance and consistency of Miniset-DenseSENet compared to the other models.

Further, we employed the MCC, sensitivity, and specificity metrics to conduct a performance evaluation of the models, as shown in Figure 2(middle, right), Figure 2(bottom, left) and Figure 2(bottom, right). The results showed that the control group performed well in terms of sensitivity and specificity. Specifically, in five experiments, the specificity of the control group for ResNet18, ResNet18+SE, and ResNet18+CBAM models reached 1, and the sensitivity for ResNet18 and Miniset-DenseSENet achieved a score of one in four out of five measurements. However, the sensitivity and specificity performance for the Concordance and Discordance classes did not meet the high standards observed in the control group and raised some stability concerns. Miniset-DenseSENet demonstrated the most superior results among them. The average sensitivity for both classes was approximately 80%, and the specificity was around 93%. In the Appendix, Figure 7 displays the confusion matrices for the six models, further validating these observations.

A comparative analysis of the six models, as observed in Table 1, shows higher computational complexity, measured in FLOPs, is generally associated with increased accuracy. However, this relationship is not linear, indicating that simply increasing computing resources does not guarantee a proportional improvement in performance [18]. The DenseNet architecture utilizing feature reuse exhibits higher parameter efficiency compared to ResNet’s residual learning method. Integrating attention mechanisms such as SE and CBAM into the traditional architectures produces mixed results. The specific reasons will be analyzed in the discussion. Miniset-DenseSENet achieves the highest accuracy among the evaluated models, indicating a positive synergy between the DenseNet architecture and the SE mechanism. In contrast, the accuracy of ResNet18+CBAM drops emphasizes that the effectiveness of the attention mechanism depends on the underlying model architecture and the specific task.

**Table 1:** Comparison of model parameters, FLOPs and average accuracy.

| Models | FLOPs | Params | AvgAccuracy |
| --- | --- | --- | --- |
| Basic CNN | 1.9G | 95232 | 63.16% |
| ResNet18 | 5.9G | 11.2M | 86.84% |
| DenseNet121 | 9.2G | 7.0M | 78.95% |
| ResNet18+SE | 5.9G | 11.2M | 81.58% |
| ResNet18+CBAM | 5.9G | 11.22M | 71.05% |
| Miniset-DenseSENet | 9.2G | 7.1M | 89.47% |

### 2.3 Grad-CAM Analysis of Correct and Incorrect Cases

We use Grad-CAM to visualize the classification rationale of the Miniset-DenseSENet trained model, exploring instances of both correct and incorrect classifications through visualization, as shown in Figure 3. The principle of creating this diagram is similar to that of confusion matrices. The horizontal axis represents the predicted categories of the images, while the vertical axis represents the true categories. From the distribution of the heatmap, it can be seen that the model correctly recognizes images along the diagonal, while misclassifications are found at other positions.

**Fig. 3:**
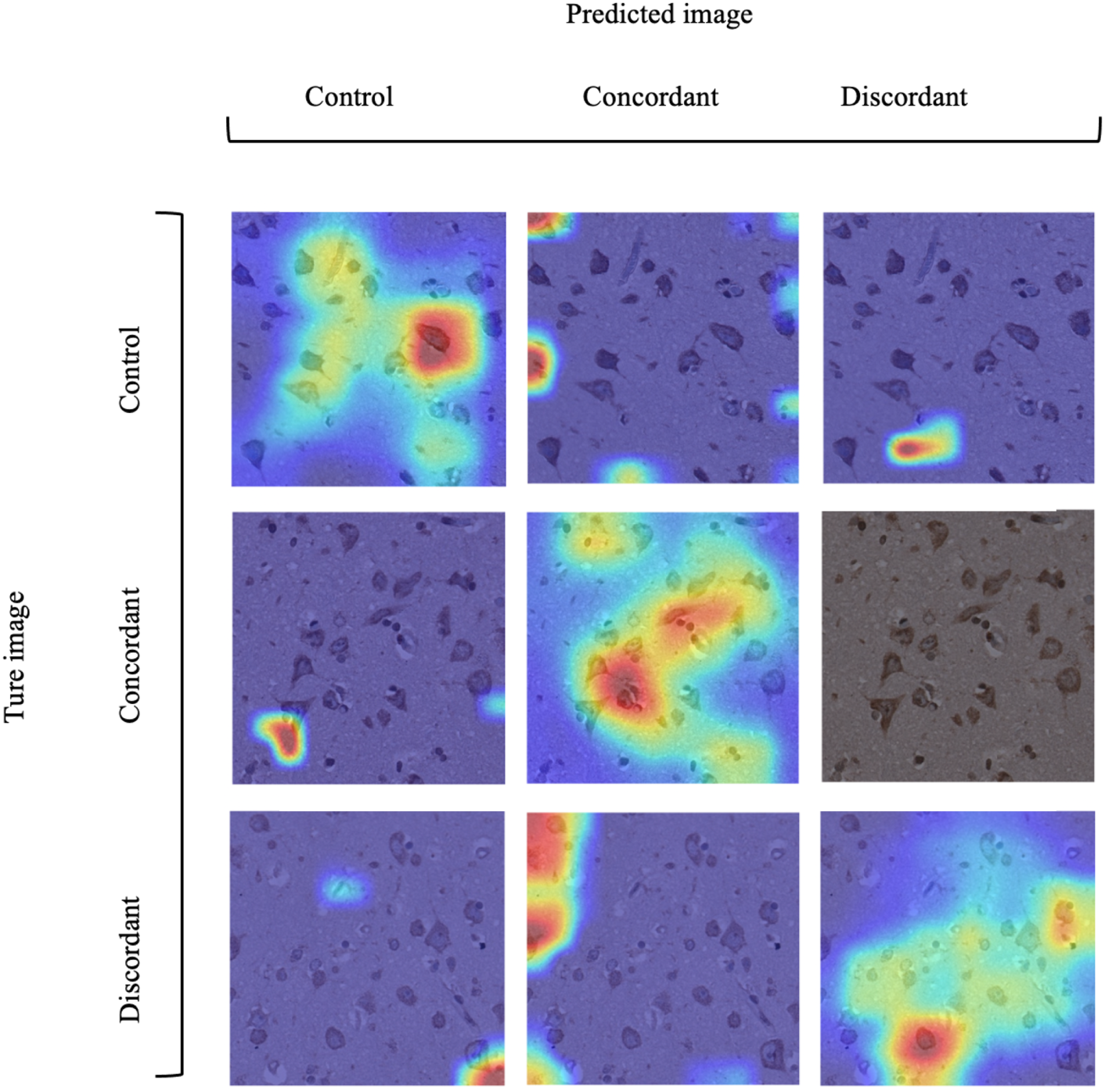
Classification Performance Analysis with Miniset-DenseSENet: Correct and Misclassified Instances. The figure presents a 3×3 matrix of class activation map (CAM) images. The columns represent the predicted labels (control, concordant, discordant), and the rows represent the actual labels. Each cell in the matrix visualizes the Grad-CAM output for a given instance, showing the areas of focus that contributed to the model’s prediction. Diagonal cells indicate correct classifications, where the predicted and actual labels match, while off-diagonal cells indicate misclassified instances, showing where the model predicted incorrectly. This visualization highlights how Miniset-DenseSENet identifies key features in different brain regions across the control, concordant, and discordant categories.

## 3 Discussion and Analysis

In this project, our decision not to employ data augmentation methods, such as colour denoising and enhancement or brightness adjustments, was motivated by the need to preserve the integrity of critical biomarkers in postmortem brain images of ALS patients. In neurodegenerative diseases, maintaining the fidelity of pathological signals is essential, especially when the underlying features of interest may be subtle or obscured by extraneous enhancements. Excessive manipulation of image characteristics can inadvertently introduce biases, causing models to focus on irrelevant or non-specific areas, thus diverging from the true disease-related features that are key to accurate diagnosis and mechanistic understanding. For ALS, where the presence of TDP-43 proteinopathy is central to the disease, ensuring that machine learning models identify and prioritise relevant features is essential.

The integration of advanced techniques such as transfer learning and attention mechanisms has significantly improved model performance compared to traditional CNN training methods (see Figure 2). Specifically, our results show that ResNet18 outperforms DenseNet121, which is noteworthy given that DenseNet models are often favoured for their deep, densely connected architectures. However, introducing the SE module into our Miniset-DenseSENet architecture shifted this balance, enabling superior performance compared to both ResNet18 and ResNet18+SE. This observation is particularly relevant in the context of machine learning applied to ALS, where data is often limited. Models like ResNet18, with fewer parameters, are advantageous when data availability is restricted, as they mitigate the risk of overfitting while still capturing disease-relevant features. However, as demonstrated by the enhanced performance of Miniset-DenseSENet, incorporating attention mechanisms through the SE module can selectively amplify critical input features in dense layers, improving the model’s capacity to generalise despite the small dataset.

The machine learning community’s ongoing work developing and refining models such as these is crucial for advancing our understanding of ALS. The pathophysiological mechanisms of ALS, particularly those involving TDP-43, are highly complex and not fully understood. Machine learning offers a unique lens through which these mechanisms can be explored, as models trained on large datasets can identify patterns and associations that may be difficult to detect through conventional analytical approaches. The integration of attention mechanisms, particularly, has shown promise in emphasising subtle but meaningful features likely to be associated with pathological changes.

However, it is important to recognise that the effectiveness of attention mechanisms is contingent upon several factors, including the complexity of the model, the nature of the dataset, and the optimisation strategy. In cases where simpler models like ResNet18 can effectively capture relevant features, further complexity may be redundant and detract from performance. This insight highlights the importance of tailoring model architecture to the specific requirements of a given task, especially in scenarios involving rare diseases like ALS, where data scarcity is often a limiting factor. The superior performance of the DenseNet121+SE model compared to ResNet18+SE may reflect the inherent compatibility of the DenseNet architecture with the SE module in this application. DenseNet’s dense connections facilitate the propagation of information across layers, which, when combined with the feature recalibration capability of the SE module, enhances the model’s ability to focus on disease-relevant areas in a more holistic manner.

This work underscores the critical role that the machine learning community plays in advancing our understanding of ALS. Optimizing model architectures and integrating innovative techniques such as attention mechanisms enhance our ability to diagnose ALS and unravel the mechanisms underlying TDP-43 aggregation. These contributions are valuable for improving clinical outcomes and deepening our biological understanding of this devastating disease. As the field progresses, we anticipate that further collaborations between machine learning researchers, neuroscientists, and clinicians will drive significant advancements in diagnosing, monitoring, and treating ALS.

## 4 Methodology

### 4.1 Image Dataset

In this project, we apply deep learning techniques to classify a dataset of 190 images provided by Gregory’s Lab at the University of Aberdeen. The dataset comprises three categories: 60 images in the concordant group, which are images of patients with TDP-43 pathology in the extramotor cortex and cognitive impairment; 60 images in the discordant group, which are images of patients with TDP-43 pathology in the extramotor cortex but without cognitive impairment. Finally, the control group consists of 70 images from individuals without ALS, characterized by the absence of TDP-43 pathology in the extramotor cortex (Figure 4).

**Fig. 4:**
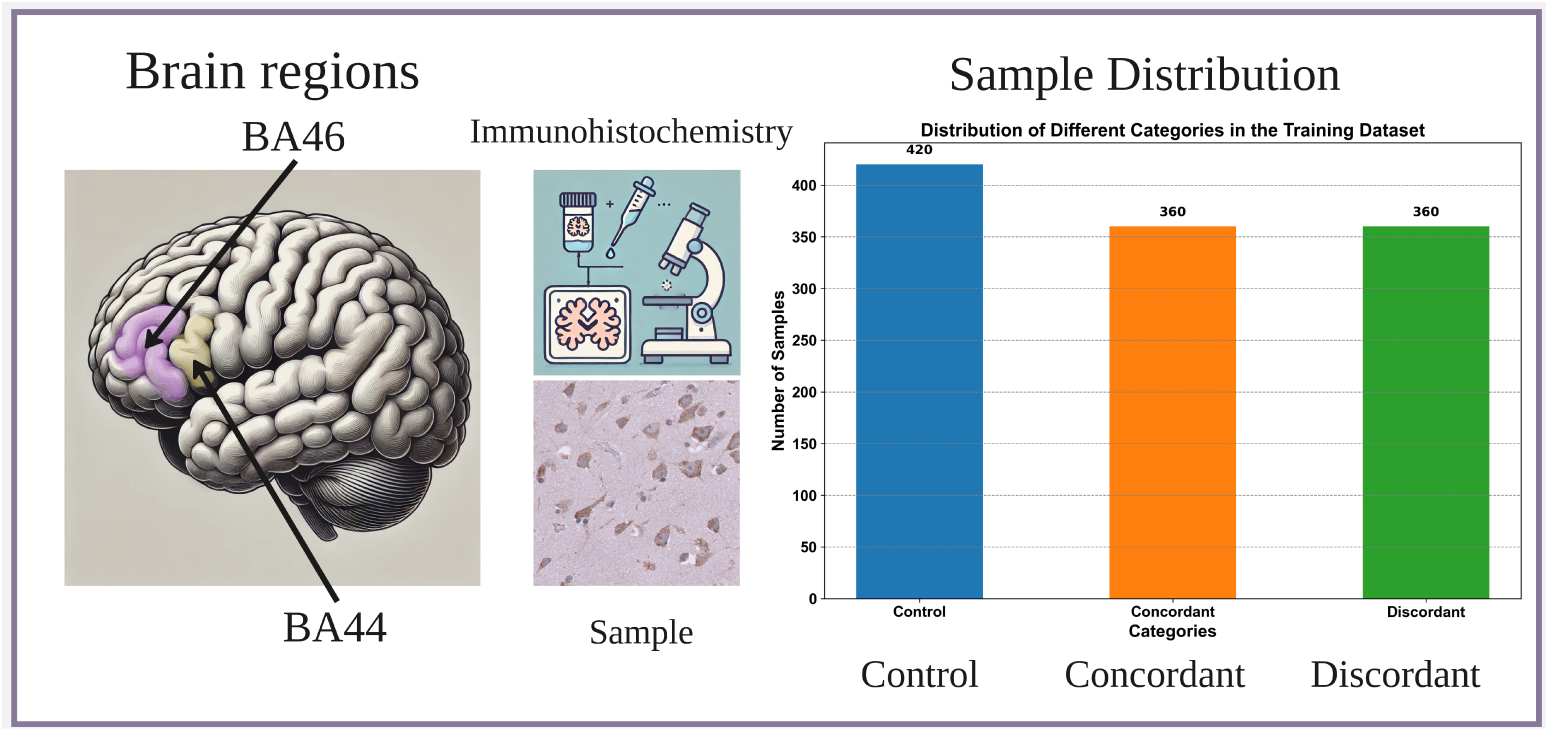
The figure provides a comprehensive overview of the dataset and its neuroanatomical focus. On the left, a brain diagram highlights the regions BA44 and BA46, which are the areas from which the brain tissue samples were taken. In the center, the top section features an icon representing the immunohistochemistry techniques used to create the brain tissue images, while below it is a sample image from the dataset. On the right, a bar chart shows the sample size distribution across three classes: control, concordant, and discordant. Concordant samples represent individuals showing signs of cognitive decline, while discordant samples display motor dysfunction without cognitive impairment. Together, these visual elements emphasize the neuroanatomical regions, image acquisition techniques, and the distribution of data across key categories in the study.

The brain tissue images were created using immunohistochemistry techniques, enabling precise staining and visualization of TDP-43 pathology in specific cortical areas. This method allows for the detection of specific proteins within the brain tissue samples, providing a detailed view of the presence and distribution of TDP-43 inclusions [4]. The images were stained with TDP-43 RNA aptamer (TDP-43APT) [19, 20], following the Standard Operating Procedure previously published [21].

The brain images were selected from two key brain regions: Brodmann area 44 (BA44) and Brodmann area 46 (BA46). BA44, located in the left inferior frontal gyrus and part of Broca’s area, is primarily associated with language production and higherorder cognitive functions [22]. BA46, situated in the dorsolateral prefrontal cortex, is involved in executive functions such as working memory, cognitive control, and decision-making [23]. These regions are of particular interest due to their involvement in both cognitive processing and motor control. This focus on BA44 and BA46 helps bridge the gap between cognitive decline and motor dysfunction, facilitating a deeper understanding of the distinct patterns observed in concordant and discordant cases.

### 4.2 Ethics Statement

All post-mortem tissue was collected with ethics approval from the East of Scotland Research Ethics Service (16/ES/0084) in line with the Human Tissue (Scotland) Act (2006). The use of post-mortem tissue for studies was reviewed and approved by the Edinburgh Brain Bank ethics committee and the Academic and Clinical Central Office for Research and Development (ACCORD) medical research ethics committee (AMREC). Clinical data were collected as part of the Scottish Motor Neurone Disease Register (SMNDR) and Care Audit Research and Evaluation for Motor Neurone Disease (CARE-MND) platform, with ethics approval from Scotland A Research Ethics Committee (10/MRE00/78 and 15/SS/0216) and have been published previously [4]).

#### 4.2.1 Dataset Division

The 190 image datasets are divided into training, validation, and test sets according to a distribution ratio of 6:2:2, as shown in Table 2. The data were split using a stratified sampling approach, ensuring that the category distribution of the original labels was preserved across the training, validation, and test sets. This method maintained class balance during the division process, which is crucial to avoid introducing bias toward the majority class, leading to reduced generalization and poorer performance on minority classes [24]. The training dataset was augmented using nine different data augmentation techniques, resulting in a total of 1,140 images in the final training set. Figure 4 (right) illustrates the categorical distribution within the training dataset.

**Table 2:** Dataset extension and division for training and testing.

| Dataset | Size | Training Size | Validation Size | Test Size |
| --- | --- | --- | --- | --- |
| Original | 190 | 114 | 38 | 38 |
| Augmented | 1140 | $(9 \times 114)$ 1026 | 38 | 38 |

### 4.3 Original Image Quality Analysis

Here, 190 images are quality-checked to ensure all training images are clear and valid. At the same time, quality inspection of the data-enhanced images can ensure the effectiveness of data enhancement. The testing of the Laplacian variance, brightness average, contrast (expressed as standard deviation), signal-to-noise ratio (SNR) and other indicators of each image allows us to analyze the quality of the dataset, following standard practices in image quality assessment [25]. In addition, we chose violin plots to show the distribution and density of these indicators, which helps us understand the fluctuations in image quality in the dataset more intuitively.

### 4.4 Data Augmentation

Training deep CNNs on small datasets and enhancing the generalization capabilities of these models is an extremely challenging task. During training, models with poor generalization tend to overfit the training data. Data augmentation is an effective strategy to solve this problem. It minimizes overfitting problems by extending the data representation to include a more comprehensive set of potential data points [12].

This project uses nine data augmentation methods, including geometric transformation and various image processing functions. Geometric transformation mainly changes the spatial layout of image content, while non-geometric transformation involves adjusting the appearance of the image, such as color, brightness, contrast, etc. The specific methods are as follows:

- Flipping: Flip the image horizontally.
- Rotating: Rotate the image 90 degrees.
- Image Cropping: Cut off a specified border width evenly from all sides of each original image to obtain a smaller central area than the original image.
- Image Scaling: Adjust the size of the image.
- Perspective Transformation: Simulates the visual effects of observing an image from different angles.
- Grayscale Conversion: Convert color images to grayscale images.
- Grayscale Image Denoising: Reduce random noise in grayscale images.
- Color Image Denoising: Reduce random noise in color images.
- Color Image Denoising: Reduce random noise in color images.
- Color Image Denoising and Enhancing Image Quality: Bilateral filtering is used to reduce noise in color images, and a binary mask is created for edge protection through the Canny edge detection [26] and expansion operations. Finally, an adaptive histogram equalization method is applied to the luminance channel of the image to improve the contrast and sharpness of the image.
- Add noise: Add random noise to the image to improve the robustness of

the model.

- Brightness Enhancement: Improve the overall brightness of the image.

Taking the original image in Figure 5 as an example, the extended images show the resulting images processed by different data augmentation methods.

**Fig. 5:**
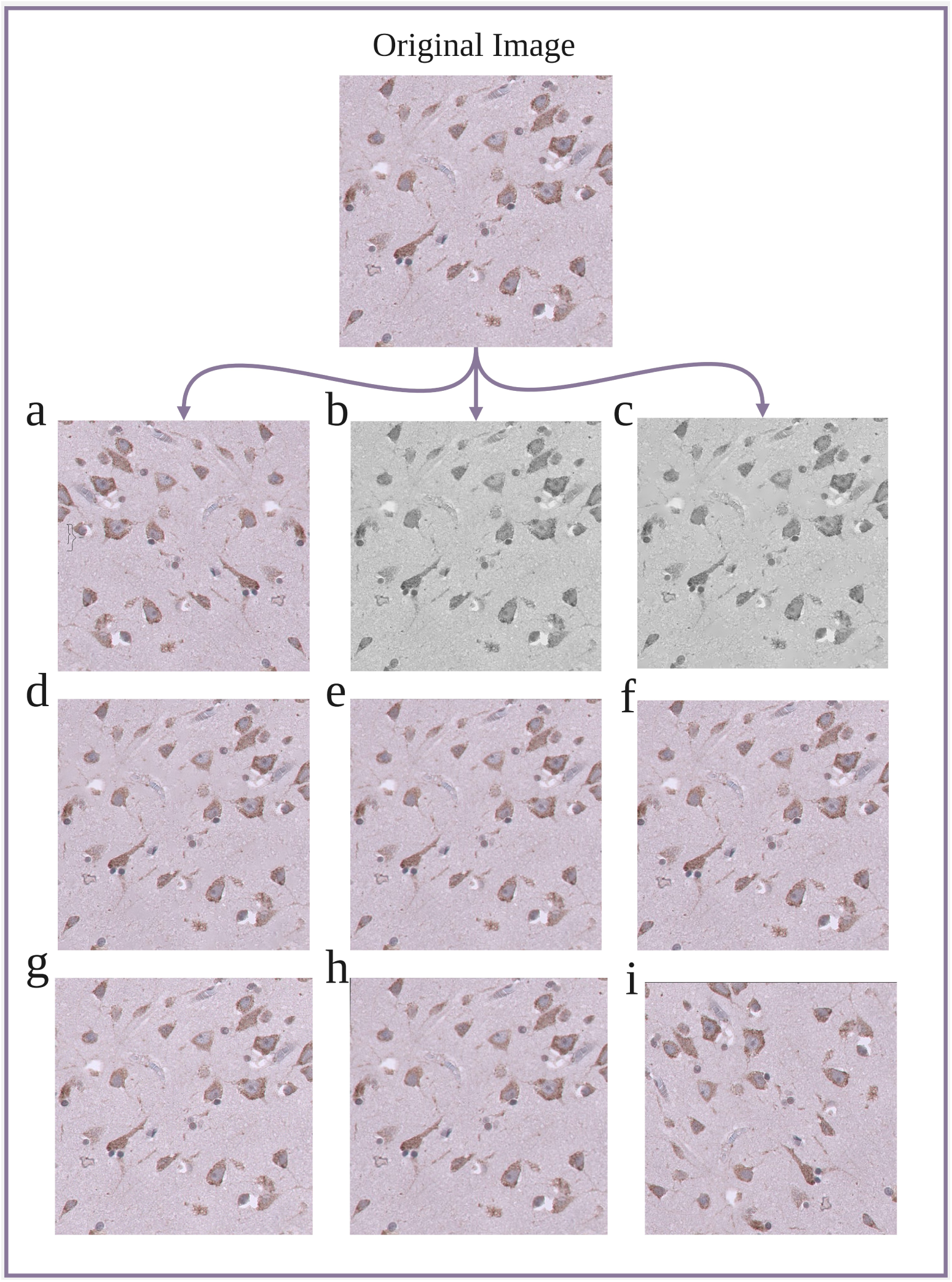
The figure above displays the original image (top), followed by nine images augmented using various techniques. The augmented images, arranged in a 3×3 matrix, are as follows: (a) Flipping, where the image is mirrored horizontally; (b) Grayscale, converting the image to grayscale; (c) GrayDenoised, applying noise reduction to the grayscale image; (d) ColorDenoising, applying noise reduction while maintaining the original color scheme; (e) Scaling, resizing the image without altering the aspect ratio; (f) Cropping, selecting and displaying only^12^a portion of the original image; (g) Adding Noise, introducing random noise to the image; (h) Perspective Transformation, altering the perspective view of the image; and (i) Rotating, rotating the image by a specified angle. These augmentations highlight the variety of transformations applied to the original data to enhance model robustness. a composite function *H_l_*, composed of convolution, batch normalization, and ReLU activation. The relationship is defined as:

### 4.5 Model Architecture

We utilized DenseNet121, a convolutional neural network known for its dense connectivity, which promotes feature reuse and mitigates the vanishing gradient problem. In this architecture, each layer is directly connected to every other layer, allowing for efficient feature transfer across layers. The output of the *l*-th layer, *x_l_*, is expressed as

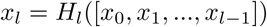

This connectivity ensures feature reuse and leads to better model regularization, which is particularly beneficial when training on smaller datasets, such as those in ALS classification tasks. DenseNet121’s inherent regularization reduces the risk of overfitting.

To enhance feature recalibration, we integrated the Squeeze-and-Excitation (SE) module into the DenseNet121 architecture. The SE module performs adaptive recalibration of channel-wise feature responses by explicitly modeling channel interdependencies. This is done through a “squeeze” operation, which aggregates spatial information into a global channel descriptor via global average pooling, followed by an “excitation” phase, which applies a Sigmoid-activated gating mechanism to selectively emphasize important features.

The recalibration process is represented as:

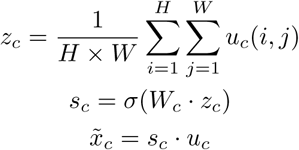

Where *u_c_*is the feature map for channel *c*, *z_c_*is the global descriptor for that channel, and *s_c_* represents the gating weights applied to recalibrate the channel’s features.

#### 4.5.1 Integration of DenseNet121 and Squeeze-and-Excitation Module

We combined DenseNet121 with the SE module, positioning the SE blocks between the dense blocks of DenseNet. This architecture, referred to as DenseNet121+SE, allows us to use DenseNet’s feature extraction capabilities while using SE modules to refine the features by suppressing irrelevant ones and enhancing critical ALS-related patterns. The overall architecture is illustrated in Figure 6, which shows the placement of SE blocks and the connection of dense blocks within the DenseNet121 backbone.

**Fig. 6:**
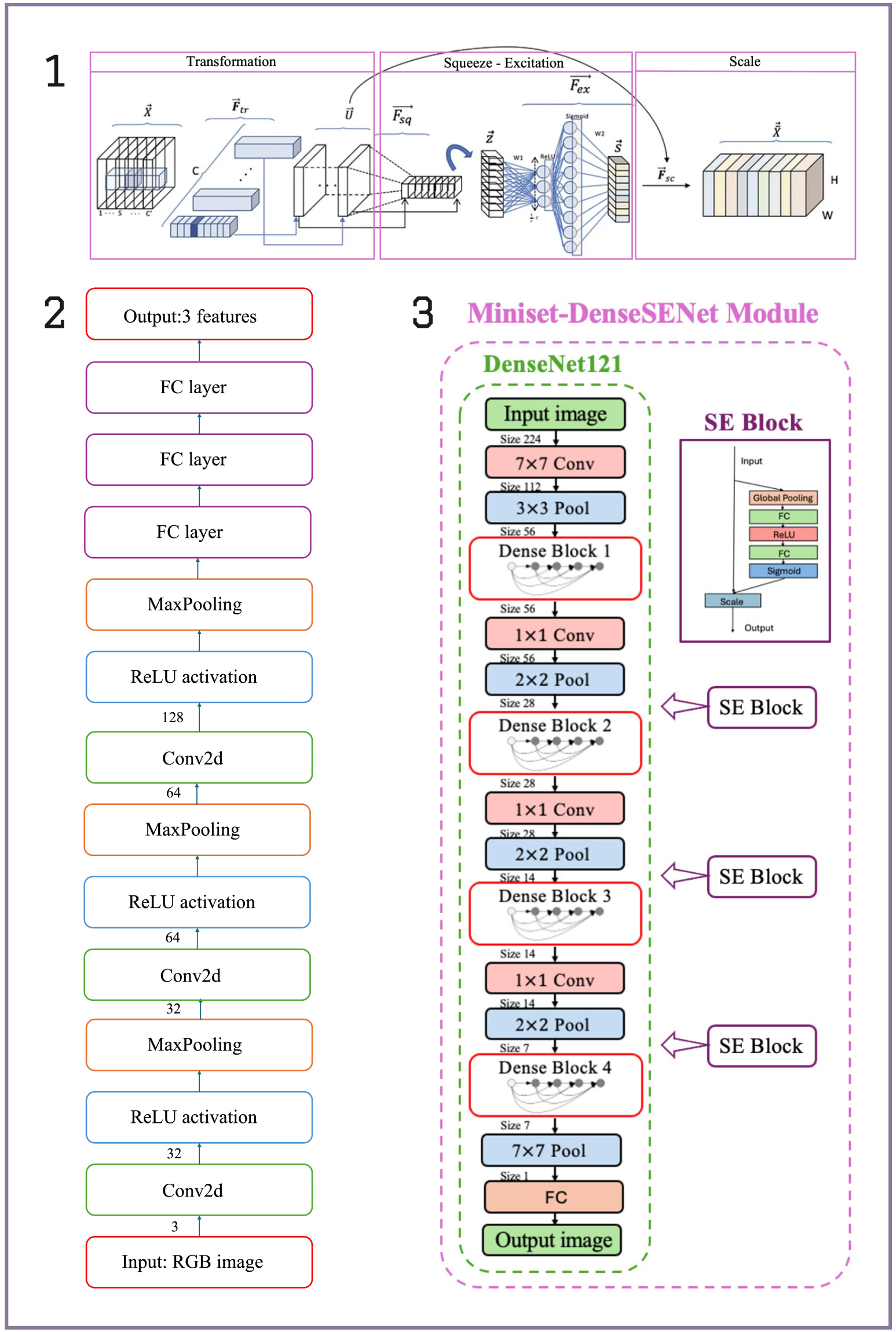
(1) The SE block enhances network performance by adaptively recalibrating channel-wise feature responses. It first uses_14_global average pooling to “squeeze” spatial information into a channel descriptor, followed by an “excitation” phase with fully connected layers that generate weights for each channel. These weights are applied to the original feature map to emphasize important features and suppress less relevant ones, improving the model’s representational capacity with minimal computational cost. (2) The model architecture of a basic CNN. (3) The model architecture of DenseNet121 and its model architecture after integrating the SE module.

**Fig. 7:**
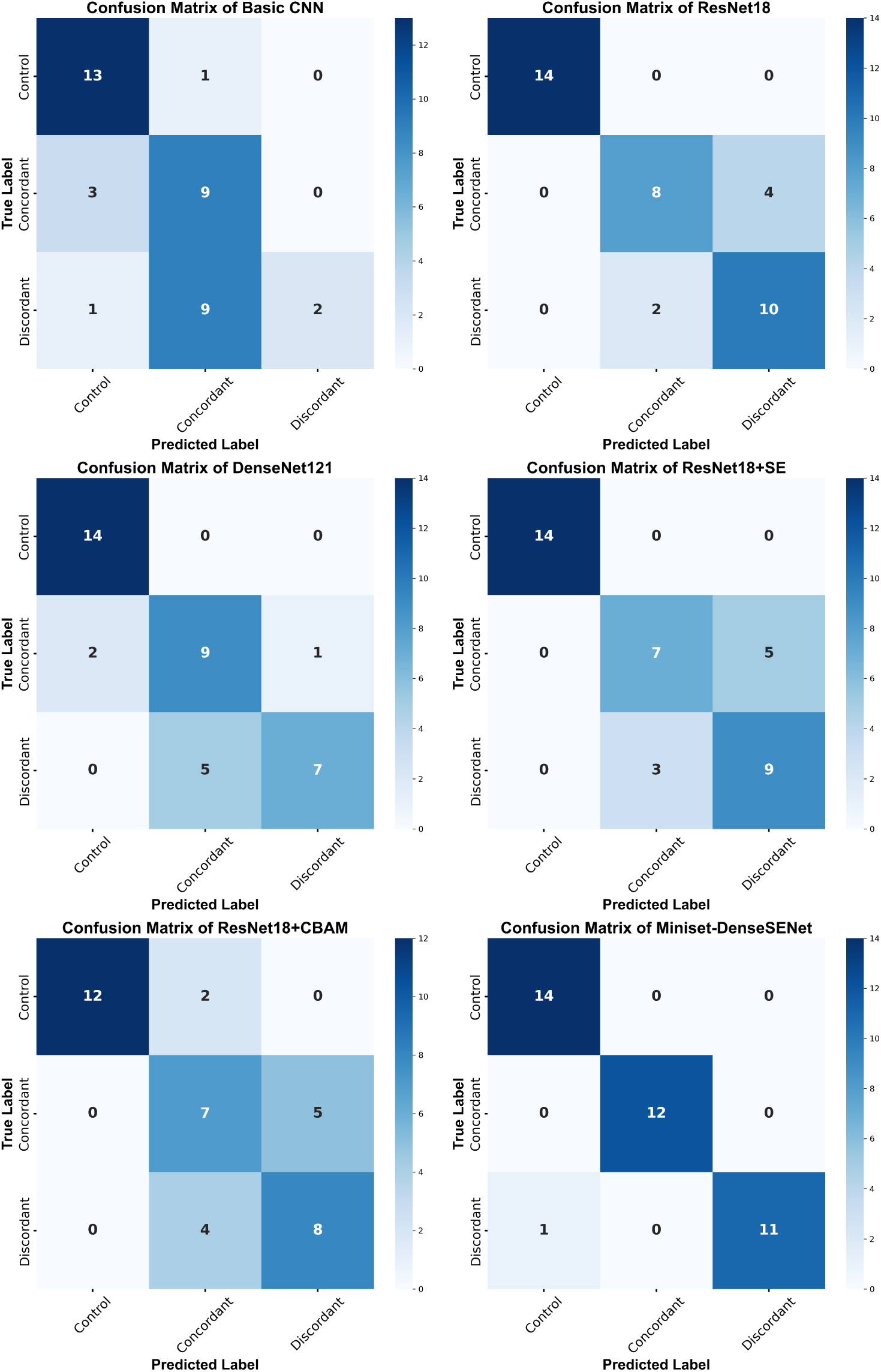
Confusion matrices generated by the Miniset-DenseSENet and the other CNN

#### 4.5.2 Training Details

The model was trained using the Adam optimizer with a learning rate of 1*e^−^*^4^ and a batch size of 16 for 50 epochs. Data augmentation techniques, such as rotation, flipping, and contrast adjustments, were applied to mitigate overfitting due to the small dataset size. The training data was split 80/20 into training and testing sets, ensuring balanced representation across ALS subtypes. We use DenseNet121 as our transfer learning model because DenseNet has efficient feature reuse and improved gradient flow.

### 4.6 Model Benchmarking

In the model training phase of this study, Python 3.8 was selected as the programming language to ensure efficient development and model implementation. The performance of Miniset-DenseSENet was compared with that of four other models: ResNet18, DenseNet121, ResNet18+SE, and ResNet18+CBAM, as well as with a simple three-layer CNN baseline model (see Figure 6(2)). These models were developed based on the foundational ResNet and DenseNet architectures, with specific enhancements through integrating SE and CBAM attention mechanisms in designated variants.

The training dataset used in this experiment includes the original dataset and data processed by nine different data augmentation techniques, with a total of 1140 images. However, given the limited scale of the available training dataset, all CNN modules in this study are pre-trained on ImageNet. When using these pre-trained models, most of the layers are frozen, with only zero to two modules in a trainable state, while the global average pooling layer and fully connected layer are set to trainable states for fine-tuning specific tasks. Here are the training strategy and freezing details:

- ResNet18: All modules of the pre-trained ResNet18 model are frozen, and only the GAP and FC are set as trainable layers.
- ResNet18+SE: All modules of the pre-trained ResNet18 model are frozen,

and the GAP, FC and SE modules are set as trainable layers.

- ResNet18+CBAM: Most modules of the pre-trained ResNet18 model are frozen, and only the last two modules, the GAP, the FC and the CBAM modules, are set as trainable layers.
- DenseNet121: All modules of the pre-trained DenseNet121 model are frozen. The GAP and the FC are set as trainable layers.
- Miniset-DenseSENet: All modules of the pre-trained DenseNet121 model are frozen, and the GAP, FC and SE modules are set as trainable layers.

The selection of hyperparameters for each model was informed by careful consideration of various factors, including limitations imposed by GPU memory, training efficiency, the risk of overfitting, and the inherent size of the models. Detailed configurations of the hyperparameters are systematically presented in Table 3.

**Table 3:** The applied hyperparameters of different CNN models.

| HPs<br>CNNs | Learning rate | Batch size | Optimizer | Epochs | Weight decay |
| --- | --- | --- | --- | --- | --- |
| ResNet18 | 0.0001 | 64 | Adam | 150 | 0.008 |
| DenseNet121 | 0.00008 | 32 | Adam | 250 | 0.08 |
| ResNet18+SE | 0.0005 | 128 | Adam | 300 | 0.0008 |
| ResNet18+CBAM | 0.0001 | 128 | Adam | 200 | 0.008 |
| Miniset-DenseSENet | 0.0001 | 32 | Adam | 200 | 0.001 |
| 3-layer CNN | 0.0001 | 64 | Adam | 300 | 0.001 |

In this project, model performance was quantified by assessing a suite of metrics, with each trained model subjected to five independent tests. The number of iterations

(epochs) of model training was set between 150 and 300. Evaluative criteria included the Matthews Correlation Coefficient (MCC) [27], accuracy, sensitivity, and specificity. MCC is a performance indicator for classification problems based on the correlation coefficient between the observed and predicted classifications. Its values range from -1 to 1, where 0 represents a random guess, 1 a perfect prediction, and -1 represents a completely inconsistent prediction. The formulas for these metrics are provided in Appendix A.3. The computational complexity, parameter efficiency, and other indicators of the models were also considered. Additionally, we performed a visual analysis of the correct and incorrect results of model classification.

### 4.7 Visualization Techniques

CNN is usually regarded as a black box model. To improve its interpretability, this study adopts the Grad-CAM technology [28]. The technique provides the model with a “visual explanation” of the decision by highlighting the areas of the image that CNN focused on when making a specific decision.

This technique relies on the gradient information of the model to generate a heatmap by analyzing the impact of specific categories on the output of the last convolutional layer. This process can be simplified by the following formula:

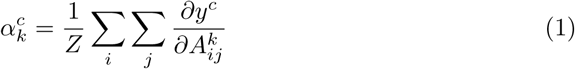

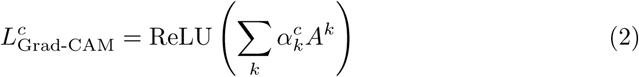

Where 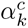 denotes the importance weights for each channel in the feature map *A^k^* with respect to class *c*. 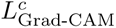 is the class-discriminative localization map for class *c*, which is a matrix of width *u* and height *v*. Before softmax transformation, *y^c^* denotes the score of category *c*, and gradient 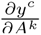 denotes the effect on the category score *y^c^* by measuring the change in feature map activation *A^k^*.

This heat map reveals which parts of the image the model values when identifying or classifying a specific class, indicating which regions contribute most to the ultimate decision. By applying Grad-CAM, researchers can visually see which part of the tissues plays a decisive role in diagnostic decisions when the model analyzes postmortem brain images of ALS patients and controls.

## 5 Conclusion and Future Work

In this study, we explored the performance of CNNs in the classification of ALS disease. We developed a model named Miniset-DenseSENet and evaluated its predictive performance using 190 autopsy brain images provided by the University of Aberdeen. Compared with standard classification models (such as ResNet18, DenseNet121, ResNet18+SE, and ResNet18+CBAM) and the baseline model (CNN), the Miniset-DenseSENet model significantly outperformed other models in classification accuracy on the small dataset.

Miniset-DenseSENet combines DenseNet121 with an SE module, leading to an increase in classification accuracy and reducing the impact of redundant features. The model’s accuracy consistently exceeds 90%, with the best performance being 97.37%. The mean value of MCC is 0.84, and the optimal result is 0.96. The sensitivity of the control category is as high as 1, and the specificity at the worst performance is as high as 0.95. The main limitation of this model is its current computational complexity. However, this limitation can be addressed through methods such as model pruning, quantization, and knowledge distillation.

The development of the Miniset-DenseSENet model holds significant importance for ALS disease. Accurate classification and early diagnosis of ALS can lead to better patient management and treatment planning. With the use of advanced neural network techniques, this model has the potential to assist medical professionals in identifying ALS with high precision, thereby contributing to improved clinical outcomes and a deeper understanding of the disease.

Considering that the proposed method is trained on a limited dataset, although the model shows high accuracy, it still has deficiencies in terms of stability. To alleviate the overfitting problem and enhance the stability of the model, it is recommended that the size of the dataset be increased. Additionally, running the model multiple times and then evaluating the results using statistical tests is suggested to determine if the observed data indicate the presence of a true effect or if it may be a random event. While the model could not accurately distinguish the location of protein density in TDP-43 aggregates, the visualization images generated by Grad-CAM have been analyzed in collaboration with our clinical team. Further refinements in visualization techniques may be needed to improve the interpretability and diagnostic utility of the model.

## Data Availability

All data produced in the present work are contained in the manuscript

https://drive.google.com/file/d/1sQwtVgqkURHpZ66bzLeh4TVnxlgniAnM/view?usp=drive_link

## 6 Competing Interests

The authors declare no competing interests.

### A Additional Material

#### A.1 Image Quality Assessment

In this study, we evaluated the quality of the dataset using several key image quality metrics to ensure that the data is suitable for further analysis and deep learning training. The following sections describe the metrics used, along with the corresponding violin plots, which confirm that the images meet the required quality standards.

##### Laplacian Variance (Sharpness)

Laplacian variance is used to assess the sharpness of each image in the dataset. A high Laplacian variance value indicates sharper images, while a lower value suggests blurriness. Sharp images are crucial for identifying fine details in brain tissue samples, which are essential for accurate feature extraction in deep learning models.

The violin plot for Laplacian variance demonstrates that the majority of images have high sharpness, with only a few outliers in the lower range. This distribution confirms that the dataset predominantly consists of clear and sharp images that are suitable for analysis.

##### Brightness Average

Brightness average measures the overall intensity of each image, ensuring that the images are neither too dark nor too bright. Proper brightness is important for distinguishing between different regions and features in the images.

The brightness average violin plot shows that the images are well-distributed around a moderate brightness level, indicating that most images have appropriate lighting conditions. This ensures that brightness will not negatively affect the quality of data processing.

##### Contrast (Standard Deviation of Intensity)

Contrast, measured as the standard deviation of pixel intensity, reflects the difference between light and dark areas in each image. Adequate contrast is necessary to highlight important features and ensure that the images contain enough detail for deep learning models to extract relevant information.

The violin plot for contrast indicates a concentrated distribution of moderate to high contrast values, suggesting that the images exhibit sufficient variability between light and dark regions. This level of contrast is optimal for deep learning tasks.

##### Signal-to-Noise Ratio

SNR assesses the level of useful information in the image relative to background noise. High SNR values indicate clearer images with less noise, which is important for accurate classification and model performance.

The SNR violin plot shows that most images have a high SNR, with only a few low-SNR outliers. This confirms that the images are generally free from excessive noise, making them appropriate for deep learning training.

Based on the analysis of these quality metrics (Laplacian variance, brightness average, contrast, and SNR), the violin plots confirm that the dataset’s image quality is acceptable. The majority of images exhibit sharpness, appropriate brightness and contrast, and low noise levels, ensuring that the dataset is valid for further data enhancement and deep learning model training.

#### A.2 Confusion Matrices

The confusion matrices presented in this section illustrate the classification performance of the models across different categories. These matrices offer insights into the accuracy of predictions for each class by displaying true positives, true negatives, false positives, and false negatives, which help in evaluating the model’s sensitivity and specificity for each class.

#### A.3 Standard Formulas for Classification Metrics

In this section, we provide the standard formulas for the classification metrics used in this study: accuracy, sensitivity, specificity, and MCC. These metrics are commonly applied in the evaluation of binary classification models [30].

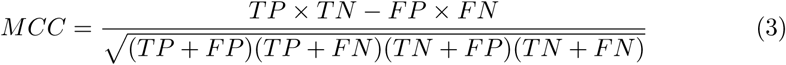

Accuracy represents the proportion of images correctly identified out of the overall number of predictions made.

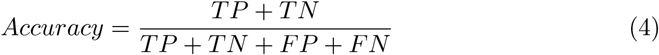

Specificity refers to the accuracy of a test in correctly recognizing individuals who are not afflicted with the disease.

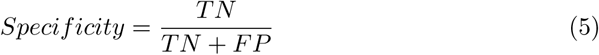

Sensitivity measures a test’s capacity to detect individuals affected by the disease accurately.

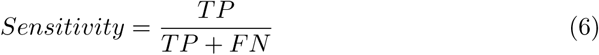

In these formulas, TP, TN, FP, and FN stand for True Positives, True Negatives, False Positives, and False Negatives, respectively.

## References

[1] Arthur, Karissa C, Calvo, Andrea, Price, T Ryan, Geiger, Joshua T, Chio, Adriano, and Traynor, Bryan J. ”Projected increase in amyotrophic lateral sclerosis from 2015 to 2040.” Nature Communications 7, no. 1 (2016): 12408.

[2] Tan, Rachel H, Ke, Yazi D, Ittner, Lars M, and Halliday, Glenda M. “ALS/FTLD: experimental models and reality.” Acta Neuropathologica 133, no. 2 (2017): 177–196.

[3] Bede, Peter. ”From qualitative radiological cues to machine learning: MRI-based diagnosis in neurodegeneration.” Future Neurology 12, no. 1 (2017): 5–8.

[4] Gregory, Jenna M, McDade, Karina, Bak, Thomas H, Pal, Suvankar, Chandran, Siddharthan, Smith, Colin, and Abrahams, Sharon. ”Executive, language and fluency dysfunction are markers of localised TDP-43 cerebral pathology in non-demented ALS.” *Journal of Neurology*, Neurosurgery & Psychiatry 91, no. 2 (2020): 149–157.

[5] Mejzini, Rita, Flynn, Loren L, Pitout, Ianthe L, Fletcher, Sue, Wilton, Steve D, and Akkari, P Anthony. ”ALS genetics, mechanisms, and therapeutics: where are we now?” Frontiers in Neuroscience 13 (2019): 497022.

[6] Caroscio, JT. ”Prognostic factors in motor neurone disease: A prospective study of longevity.” Research Progress in Motor Neuron Disease, 34–43. Pitman Books Limited, 1984.

[7] Louwerse, ES, Visser, CE, Bossuyt, PMM, Weverling, GJ, and others. ”Amyotrophic lateral sclerosis: mortality risk during the course of the disease and prognostic factors.” Journal of the Neurological Sciences 152 (1997): s10–s17.

[8] Hussain, Mahbub, Bird, Jordan J, and Faria, Diego R. ”A study on CNN transfer learning for image classification.” In Advances in Computational Intelligence Systems: Contributions Presented at the 18th UK Workshop on Computational Intelligence, September 5-7, 2018, Nottingham, UK, 191–202. Springer, 2019.

[9] Alzahrani, A Khuzaim, Alsheikhy, Ahmed A, Shawly, Tawfeeq, Azzahrani, Ahmad S, and AbuEid, Aws I. ”Amyotrophic lateral sclerosis prediction framework using a multi-level encoders-decoders-based ensemble architecture technology.” Journal of King Saud University-Computer and Information Sciences, 101960. Elsevier, 2024.

[10] Atmaramani, Rahul, Dreossi, Tommaso, Ford, Kevin, Gan, Lin, Mitchell, Jana, Tu, Shengjiang, Velayutham, Jeevaa, Zeng, Haoyang, Chickering, Michael, and Soare, Tom. ”Deep Learning Analysis on Images of iPSC-derived Motor Neurons Carrying fALS-genetics Reveals Disease-Relevant Phenotypes.” bioRxiv, 2024-01. Cold Spring Harbor Laboratory, 2024.

[11] Han, Dongmei, Liu, Qigang, and Fan, Weiguo. ”A new image classification method using CNN transfer learning and web data augmentation.” Expert Systems with Applications 95 (2018): 43–56.

[12] Shorten, Connor, and Khoshgoftaar, Taghi M. ”A survey on image data augmentation for deep learning.” Journal of Big Data 6, no. 1 (2019): 1–48.

[13] Chlap, Phillip, Min, Hang, Vandenberg, Nym, Dowling, Jason, Holloway, Lois, and Haworth, Annette. ”A review of medical image data augmentation techniques for deep learning applications.” Journal of Medical Imaging and Radiation Oncology 65, no. 5 (2021): 545–563.

[14] Weiss, Karl, Khoshgoftaar, Taghi M, and Wang, DingDing. ”A survey of transfer learning.” Journal of Big Data 3 (2016): 1–40.

[15] Raghu, Maithra, Zhang, Chiyuan, Kleinberg, Jon, and Bengio, Samy. ”Transfusion: Understanding transfer learning for medical imaging.” Advances in Neural Information Processing Systems 32 (2019).

[16] Chen, Leiyu, Li, Shaobo, Bai, Qiang, Yang, Jing, Jiang, Sanlong, and Miao, Yanming. ”Review of image classification algorithms based on convolutional neural networks.” Remote Sensing 13, no. 22 (2021): 4712.

[17] Niu, Zhaoyang, Zhong, Guoqiang, and Yu, Hui. ”A review on the attention mechanism of deep learning.” Neurocomputing 452 (2021): 48–62.

[18] Li, Wei, Zhu, Xiatian, and Gong, Shaogang. ”Harmonious attention network for person re-identification.” In Proceedings of the IEEE conference on computer vision and pattern recognition, 2285–2294. 2018.

[19] Spence, Holly, Waldron, Fergal M, Saleeb, Rebecca S, Brown, Anna-Leigh, Rifai, Olivia M, Gilodi, Martina, Read, Fiona, Roberts, Kristine, Milne, Gillian, and Wilkinson, Debbie. ”RNA aptamer reveals nuclear TDP-43 pathology is an early aggregation event that coincides with STMN-2 cryptic splicing and precedes clinical manifestation in ALS.” Acta Neuropathologica 147, no. 1 (2024): 50.

[20] Zacco, Elsa, Kantelberg, Owen, Milanetti, Edoardo, Armaos, Alexandros, Panei, Francesco Paolo, Gregory, Jenna, Jeacock, Kiani, Clarke, David J, Chandran, Siddharthan, and Ruocco, Giancarlo. ”Probing TDP-43 condensation using an in silico designed aptamer.” Nature Communications 13, no. 1 (2022): 3306.

[21] Waldron, Fergal M, Spence, Holly, and Gregory, Jenna. ”TDP-43 RNA aptamer staining to detect pathological TDP-43 in FFPE human tissue, as described in Spence and Waldron, et al., 2024 (Acta Neuropathologica): A SOP and tick-sheet. v2.” 2024.

[22] Amunts, Katrin, and Zilles, Karl. ”Architecture and organizational principles of Broca’s region.” Brain and Language 123, no. 2 (2012): 125–135.

[23] Fuster, Joaqúın M. ”The prefrontal cortex—an update: Time is of the essence.” Neuron 30, no. 2 (2001): 319–333.

[24] Johnson, Justin M, and Khoshgoftaar, Taghi M. ”Survey on deep learning with class imbalance.” Journal of Big Data 6, no. 1 (2019): 1–54.

[25] Wang, Zhou, and Bovik, Alan C. ”Modern image quality assessment.” Synthesis Lectures on Image, Video, and Multimedia Processing 2, no. 1 (2006): 1–156.

[26] Canny, John. ”A computational approach to edge detection.” IEEE Transactions on Pattern Analysis and Machine Intelligence, no. 6 (1986): 679–698.

[27] Chicco, Davide, and Jurman, Giuseppe. ”The advantages of the Matthews correlation coefficient (MCC) over F1 score and accuracy in binary classification evaluation.” BMC Genomics 21 (2020): 1–13.

[28] Selvaraju, Ramprasaath R, Cogswell, Michael, Das, Abhishek, Vedantam, Ramakrishna, Parikh, Devi, and Batra, Dhruv. ”Grad-CAM: Visual explanations from deep networks via gradient-based localization.” In Proceedings of the IEEE international conference on computer vision, 618–626. 2017.

[29] Olivia M. Rifai, James Longden, Judi O’Shaughnessy, Michael D.E. Sewell, Judith Pate, Karina McDade, Michael J.D. Daniels, Sharon Abrahams, Siddharthan Chandran, Barry W. McColl, et al., ”Random forest modelling demonstrates microglial and protein misfolding features to be key phenotypic markers in C9orf72-ALS”, The Journal of Pathology, vol. 258, no. 4, pp. 366–381, 2022.

[30] Author, A., Another, B., and Contributor, C. (2021). Title of the Study on Classification Metrics. Journal Name, Volume(Issue), Pages.

